# Application of the Muse portable EEG system to aid in rapid diagnosis of stroke

**DOI:** 10.1101/2020.06.01.20119586

**Authors:** Cassandra M. Wilkinson, Jennifer I. Burrell, Jonathan W. P. Kuziek, Sibi Thirunavukkarasu, Brian H. Buck, Kyle E. Mathewson

## Abstract

**Objective:** In this pilot study, we investigated using portable electroencephalography (EEG) as a potential prehospital stroke diagnostic method.

**Methods:** We used a portable EEG system to record data from 25 participants, 16 had acute ischemic stroke events, and compared the results of age-matched controls that included stroke mimics. Delta/alpha ratio (DAR), (delta+theta)/(alpha+beta) ratio (DBATR) and pairwise-derived Brain Symmetry Index (pdBSI) were investigated, as well as accelerometer and gyroscope trends. We then made classification trees using TreeBagger to distinguish between different subgroups.

**Results:** DAR and DBATR showed an increase in ischemic stroke patients that correlates with stroke severity (p<0.01, partial η^2^ = 0.293; p<0.01, partial η^2^ = 0.234). pdBSI decreased in low frequencies and increased in high frequencies in patients who had a stroke (p<0.05, partial η^2^ = 0. 177). All quantitative EEG measures were significant between stroke patients and controls. Using classification trees, we were able to distinguish between subgroups of stroke patients and controls.

**Conclusions:** There are significant differences in DAR, DBATR, and pdBSI between patients with ischemic stroke when compared to controls; results relate to severity.

**Significance:** With significant differences between patients with strokes and controls, we have shown the feasibility and utility for the Muse™ EEG system to aid in patient triage and diagnosis as an early detection tool.

## Introduction

Stroke is a devastating disease and the leading cause of disability in Canada.^1^ For patients with ischemic stroke, early reperfusion with either thrombolysis or endovascular devices is the most effective means of saving brain tissue and improving recovery. Stroke diagnosis begins in the prehospital setting. Emergency medical services (EMS) crews examine patients with stroke symptoms using standardized scales that are designed to separate stroke from other conditions that mimic stroke before making transport decisions regarding the destination hospital.^2^ Ensuring an accurate prehospital stroke diagnosis is essential to ensure that stroke patients are efficiently transported to hospitals that offer emergent reperfusion therapies while avoiding unnecessary transport of stroke mimics. Strokes scales assign points to clinical deficits based on an abbreviated version of the neurological exam. These prehospital stroke scales miss up to 30% of acute strokes^3^ and often miss stroke from large vessel occlusions (LVO) that benefit from endovascular thrombectomy.^4^ There remains a pressing need for rapid, cost-effective technologies to improve the prehospital diagnosis of stroke.^5^

Electroencephalography (EEG), or recording the brain’s electrical activity, has been explored as a diagnostic and prognostic tool in stroke.^6,7^ When cerebral blood flow (CBF) drops during ischemia, abnormalities such as decreases in fast frequencies are observed.^8^ Abnormalities, including slowing (increased delta to alpha ratio, DAR or increased (delta+theta)/(alpha+beta) ratio, DTABR) and changes in brain symmetry, are well established in the acute phase of stroke.^9,10^ The power spectrum begins to return to normal in the first three months after stroke, but quantitative EEG (qEEG) can still show abnormalities, such as impaired symmetry, that persist past the point of good motor recovery.^11^ Past studies have shown that qEEG indices such as DAR can be used for both diagnosis of ischemic stroke^6,8,12,13^ and prediction of clinical outcomes.^7,14-18^

Most strokes are unilateral, and thus deficits (e.g., motor impairments) are lateralized as well.^19^ Therefore, brain symmetry is another quantitative EEG measure that has been used to distinguish stroke from control populations.^7,8,12-14,18^ The brain symmetry index was first used during carotid surgery as a reliable method to detect early brain ischemia.^20^ Derived from the brain symmetry index, the pairwise-derived brain symmetry index (pdBSI) calculates asymmetry by calculating the absolute value of the difference in power at each pair of electrodes.^14^ Therefore, a larger pdBSI represents more asymmetry between the hemispheres. Higher pdBSI is often associated with worse outcome.^7,10,21^ However, it has also been shown that pdBSI may vary based on stroke location, with cortical strokes having more symmetrical activity compared to subcortical strokes.^12^

These qEEG measures of post-stroke brain activity have also been found in animal models. After a transient middle cerebral artery occlusion (MCAO) in rats, DAR increases in the acute phase and begins to recover in the chronic phase (7 days post-stroke).^22^ Behavioural deficits correlated strongly with DAR in this rodent model. Additionally, although DAR recovers spontaneously after stroke, DAR recovers more efficiently in animals that underwent reperfusion after MCAO.^23^ compared to those given a permanent MCAO. In this large stroke model, a decrease in brain symmetry is observed, primarily in the regions affected by the stroke. These animal studies are beneficial, as they both validate findings seen clinically and provide continuous data over a longer period, giving insight into the electroencephalographic changes over the course of stroke recovery.

Despite successes finding predictive quantitative EEG measures, barriers exist that currently prevent EEG from being routinely used clinically as a diagnostic or monitoring device. Commonly, the set-up and recording of typical EEG devices can take valuable time, with many set-ups in past studies using nineteen or more electrodes.^16^ In an acute and time-sensitive setting, such as an ambulance or an emergency room, the routine set-up of these systems would not be feasible. Further, access to both EEG devices and expertise can be limited. Demand exists for an EEG device that is cost-effective, simple to set-up, and easy to interpret. Advances in technology have led to consumer wireless EEG systems that meet these requirements. The Muse™ by InteraXon Inc. is one type of wearable consumer EEG system that is commercially available and has been compared to medical EEG devices.^24^ Ratti et al. found the Muse™ required significantly less time to set up than the medical system, although this was accompanied with increased test-retest variability, lower data quality, and more artifacts due to eye blinks and muscle movement.^25^ Wearable EEG has been successful in diagnostic feasibility for other disorders, such as epilepsy.^26^ Recently a pilot study was published that suggested stroke patients could be distinguished from healthy controls using the Muse™ headband due to significantly increased brain asymmetry in stroke patients.^27^ However, this study used healthy volunteers as controls rather than stroke mimics, and it is possible that stroke mimics also present with EEG abnormalities. Further, they did not determine the sensitivity of brain asymmetry as a measure to predict stroke occurrence, so further research is needed. If the data acquired using the Muse™ has enough sensitivity to distinguish stroke patients from controls, this inexpensive and portable device could be easily implemented in ambulances for improved diagnosis and triage to determine the most appropriate hospital (e.g., primary stroke centre).

The main aim of this study was to examine the feasibility of using the Muse™ in a subacute stroke population to distinguish ischemic stroke patients from a control group that included patients who presented with suspected stroke (i.e., “stroke mimics”). Further, we determined if EEG data can detect differences in stroke severity. We used classification trees to determine the most predictive combination of EEG and other physiological parameters, and calculated the sensitivity and specificity of these models in predicting stroke presence and stroke severity.

## Methods

### Participants

Twenty-one patients presenting with stroke deficits were prospectively recruited from the stroke unit at the University of Alberta Hospital (Edmonton, Alberta). The diagnosis of ischemic stroke and stroke severity was confirmed by a neurologist blind to the EEG results, based on a review of the clinical information and neuroimaging that included multimodal CT and diffusion-weighted MRI. Stroke severity was graded based on the National Institutes of Health stroke scale (NIHSS) and size of infarct. Of the 21 patients, five had negative diffusion-weighted MRI and were included in the control group. The patients’ family members were recruited as an age-matched control, bringing our total sample to 25 participants (16 stroke patients and nine controls). Due to connectivity issues, two additional sessions were excluded from the analysis. Demographic information for each participant is available in Table 1. Participants were between the ages of 19 and 91 (mean age of 66). The mean recording time was 3.85 days post-stroke onset, and the average NIHSS was 7.7. Table 1 provides a summary of demographic information sorted by the severity of the stroke event.

**Table 1:** Participant demographics and characteristics and summary by severity grouping

| **Subject Number** | **Stroke or Control** | **Severity** | **Stroke Location** | **Lateralization** | **Time from Stroke Onset** | **NIHSS** | **Gender** | **Age** |
| --- | --- | --- | --- | --- | --- | --- | --- | --- |
| 1 | Control | Control | - | - | - | - | M | 91 |
| 2 | Stroke | Small | Cortical | Left | 6 days | 0 | M | 87 |
| 3 | Stroke | Moderate | Cortical | Left | 7 days | 8 | F | 61 |
| 4 | Stroke | Moderate | Cortical | Left | 1 day | 12 | M | 65 |
| 5 | Stroke | Moderate | Subcortical | Left | 3 days | 16 | F | 83 |
| 6 | Stroke | Small | Subcortical | Right | 3 days | 0 | F | 19 |
| 7 | Stroke | Moderate | Cortical | Right | 2 days | 1 | F | 71 |
| 8 | Stroke | Moderate | Cortical | Left | 4 days | 14 | M | 71 |
| 9 | Stroke | Small | Cortical | Left | 1 day | 8 | F | 86 |
| 10 | Stroke | Moderate | Cortical | Left | 0 days | 10 | M | 85 |
| 11 | Stroke | Moderate | Cortical | Left | 8 days | 15 | M | 37 |
| 12 | Stroke | Large | Subcortical | Right | 16 days | 10 | M | 87 |
| 13 | Control | Control | - | - | - | - | M | 66 |
| 14 | Stroke | Large | Cortical | Left | 2 days | 14 | F | 53 |
| 15 | Control | Control | - | - | - | - | M | 53 |
| 16 | Stroke | Small | Cortical | Left | 3 days | 1 | M | 66 |
| 17 | Control | Control | - | - | - | - | F | 64 |
| 18 | Control | Control | - | - | - | - | F | 81 |
| 19 | Stroke | Moderate | Cortical | Left | 2 days | 2 | M | 75 |
| 20 | Control | Control | - | - | - | - | M | 56 |
| 21 | Stroke | Moderate | Cortical | Left | 0 days | 10 | M | 87 |
| 22 | Control | Control | - | - | - | - | M | 59 |
| 23 | Control | Control | - | - | - | - | F | 48 |
| 24 | Stroke | Large | Cortical | Right | 6 Days | 10 | F | 72 |
| 25 | Control | Control | - | - | - | - | M | 29 |
| **Summary Data** |  |  |  |  |  |  |  |  |
|  | - | Control (9) |  | N/A | N/A | N/A | 67% Male | 60.8 |
|  | - | Small (5) |  | 75% Left | 3.25 days | 2.25 | 33% Male | 64.5 |
|  | - | Moderate (9) |  | 89% Left | 3 days | 9.78 | 67% Male | 70.6 |
|  | - | Large (3) |  | 67% Left | 8 days | 11.33 | 50% Male | 70.7 |

#### Patient Consent

Written informed consent was obtained from participants or their legal guardians if they could not consent prior to participating. The Human Research Ethics Office, University of Alberta, Canada, approved ethics for this study.

#### Data Availability Statement

Anonymized data will be shared upon request from qualified investigators.

### Procedure

Each experiment began by having the procedure explained to the participant or their guardian, then receiving informed consent. The participant’s forehead and earlobes were cleaned with NuPrep, an exfoliating gel, then cleaned with alcohol swabs to ensure a stable connection. The Muse™ (InteraXon Inc., Toronto, ON) was cleaned with alcohol swabs before and after each recording. EEG recordings consisted of two sessions, three minutes in an eyes-open resting state and another three minutes in an eyes-closed resting state. Eyes-open resting-state consisted of the patient concentrating at a fixation cross centred on a 13” MacBook Air Laptop running macOS High Sierra version 10.13.3. Recordings were taken with a Muse™ headband that had its hard-plastic exterior removed and replaced with a soft headband, secured with an adjustable elastic cord strap, for improved electrode connection. The Muse™ was further modified as the ear electrodes were replaced with clip-on electrodes to ensure a better connection. The headband sat roughly 2.5cm above the eyebrows. All electrodes were silver chloride coated and connected with electrolyte gel to further increase connectivity.

### EEG Analysis

The Muse™ has a 256 Hz sampling frequency. The Muse™ consists of seven electrodes, four electrodes located at A7, A8, T9 and T10, a ground electrode located at Fpz, and two reference electrodes to the left and right of the ground. These electrodes roughly correspond to the international 10-20 electrode system. The Muse™ has an on-board digital signal processing module that performs noise filtering. Data was recorded using MuseLSL.^28^ Data was then transmitted at 10 Hz via a Silicon Labs Bluetooth Module (Model: BLED112), for further analysis in MATLAB.

Oscillations in the data were estimated using a wavelet transform with a wavenumber of 15. Power was calculated for frequencies between 0.5 and 31 in steps of 0.1. Wavelet code was used from the BOSC (Better OSCillation detection) toolbox.^29^ Only data from the eyes closed portion of the recording was used for further analysis, as some patients were unable to keep their eyes open for the three-minute recording window. Only TP9 and TP10 electrodes were used for qEEG measures, as some patients had connectivity issues with the frontal electrodes. The entire three-minute data segment was averaged together to create a single power spectrum at each electrode for each participant. Then the global asymmetry was calculated using homologous electrode pairs in the 1-30 Hz range. We used the calculation from Sheorajpanday et al. 2011, which defined pdBSI as:

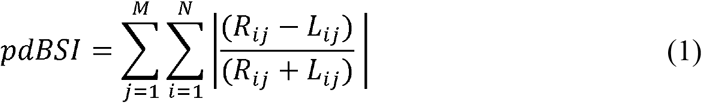

with R_ij_ and L_ij_ being the power spectral density of the signals for each electrode pairing (with i = 1, 2,…, M) at each frequency (with j = 1, 2,…, N).^10^ DAR was calculated as the sum of power of delta (1–3 Hz) frequencies divided by the sum of power of alpha (8-13 Hz) frequencies. We calculated DTABR as the sum of power of delta (1-3 Hz) and theta (4–7 Hz) frequencies divided by the sum of power of alpha (8–13 Hz) and beta (14-20 Hz) frequencies. In addition to these band-based measures of oscillatory power, we also analyzed the spectra using Fitting Oscillations & One-Over F (fooof),^30^ to determine the slope and intercept of the aperiodic component to see if these differed between groups. This analysis was performed using version 0. 1.1 of the FOOOF MATLAB wrapper with the following settings: peak width limits = [0.5,12]; max number of peaks = Inf; minimum peak amplitude = 0.0; peak threshold = 2.0; background mode = fixed; verbose = true; frequency range = 0.5-30 Hz. Finally, using the onboard gyroscope and accelerometer, we calculated both the standard deviations and the root mean square (RMS) of the head movement over time to identify the differences in variability of movement across the X, Y, and Z movement planes.

Using data calculated in the previous analyses (Table 2), TreeBagger creates classification trees by utilizing the inputted data as base learners, then applying a bootstrapping algorithm to produce N number of trees (N = 200), then combines multiple trees to produce one final tree thus reducing the effect of overfitting and improving generalization. We created three trees to predict the severity of strokes, if strokes had occurred, and the final tree determined whether the participant fell into a higher-risk category. This final model classified moderate and large strokes from small strokes and controls.

**Table 2:**
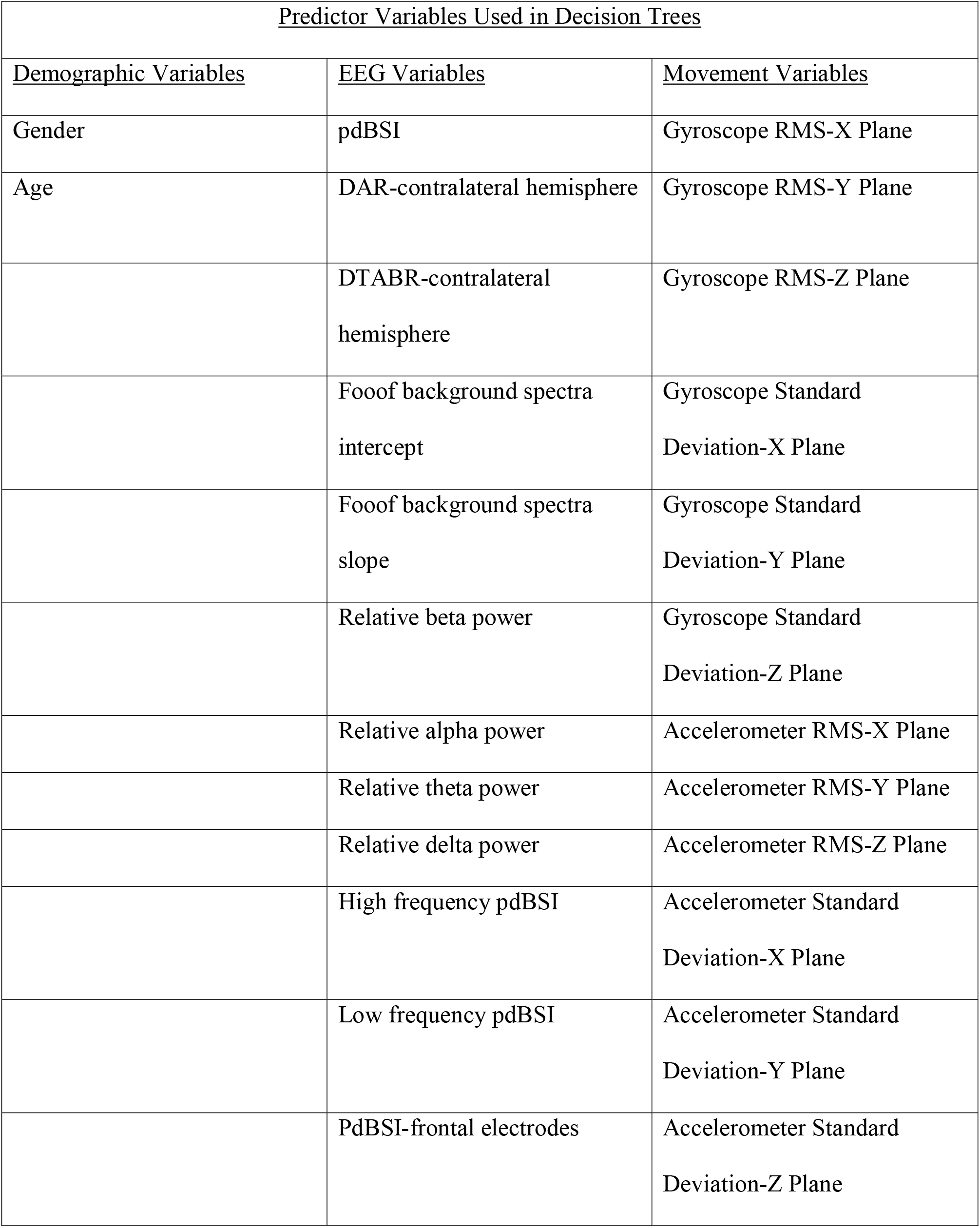
List of predictor variables used in making decision trees

### Statistical Analysis

All statistical analyses were completed in MATLAB with the significance threshold of α = 0.05. We used a one-way ANOVA to test for group differences in pdBSI. We ran 2-way ANOVA tests on the outcomes of the DAR, DTABR, gyroscope, and accelerometer data with factors of severity and hemisphere. Post-hoc group comparisons were done using the Tukey-Kramer correction. All data are presented as mean ± standard error of the mean (SEM). We calculated accuracy (ACC), sensitivity (SE) and specificity (SP) as a comparison of each classification tree output and the outcome of clinical diagnoses (equations 2-4, respectively). These calculations used the number of true positives (TP), false positives (FP), false negatives (FN), and true negatives (TN).^31^

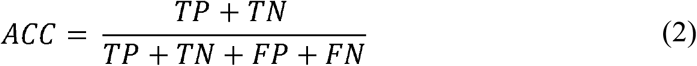

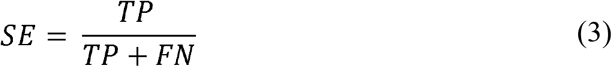

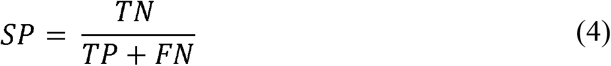

## Results

The analysis of pdBSI showed significant differences between patients with strokes and the healthy controls (p < 0.05, F = 4.96, df = 1, 23, partial η2 = 0.177). Overall, brain activity was more symmetrical in stroke patients (Figure 1A). Changes in pdBSI were dependent on the frequency range, as seen in Figure 1C. In stroke patients, brain symmetry decreased at lower frequencies and increased at higher frequencies, whereas the opposite is true for the controls (Figure 2). pdBSI also showed a trending severity effect, although insignificant as indicated by post-hoc tests (Figure 1B). This severity effect was frequency-dependent and was driven by the beta frequency range, while an opposing effect is seen in the delta range as expected from the spectra in Figure 2.

**Figure 1.**
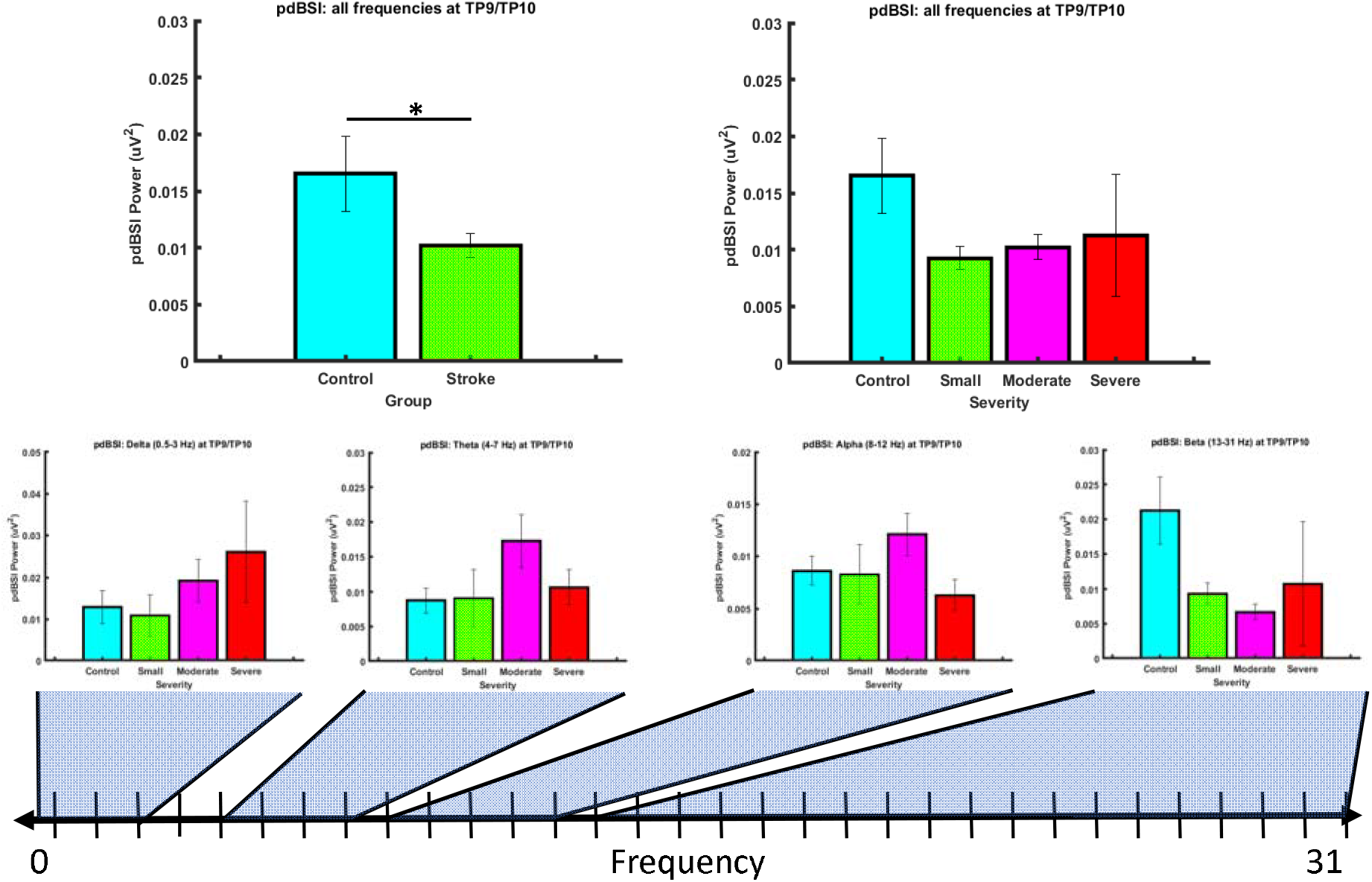
Pairwise Derived Brain Symmetry Index by group and frequency. TP9 and TP10, (B) pdBSI by severity across all frequencies, (C) pdBSI by severity for each frequency bin. Standard error was calculated for each bar. * significance at p<0.050. Brain activity is more symmetrical after stroke. Average pdBSI does not readily distinguish between stroke severities. Data is presented as mean ± SEM.

**Figure 2.**
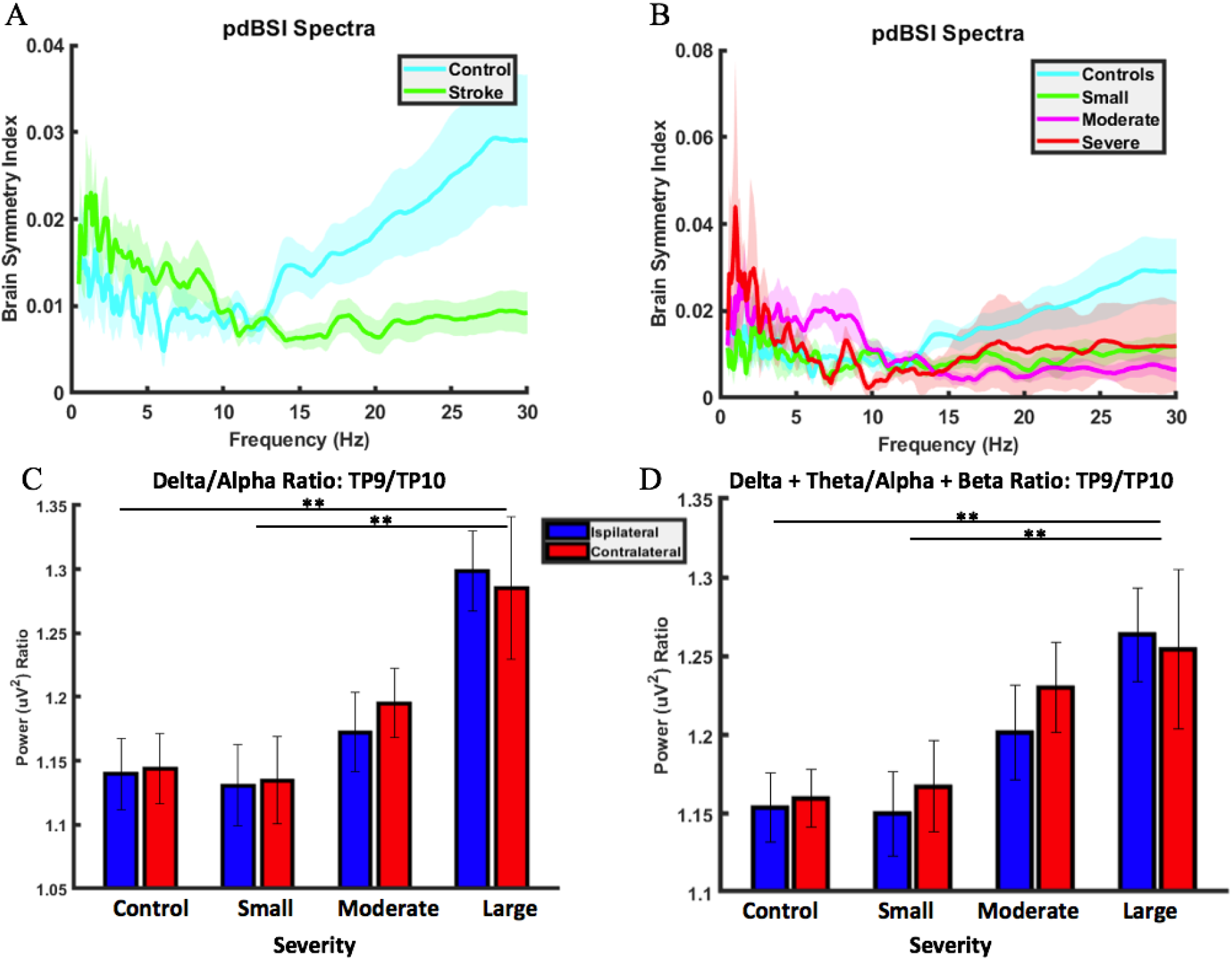
Pairwise Derived Brain Symmetry Index across frequency by stroke severity at electrodes TP9 and TP10, DAR across severities, and DTABR across severities. ** significance at p=0.01. In stroke patients, pdBSI increases at low frequencies (A). However, above ~ 10 Hz, pdBSI decreases in stroke patients. When stroke patients are grouped by severity (B), it is difficult to differentiate severity by pdBSI at higher frequencies, largely due to high variability in the severe stroke group. DAR increases as stroke severity increases in moderate and severe strokes (C). We do not observe any clear hemispheric differences in DAR. DTABR increases with increasing severity (D). No clear hemispheric differences have been observed, but in small and moderate stroke there is a trend for increased slowing on the injured hemisphere. Data are presented as mean ± SEM.

We calculated DAR as seen in Figure 2C; we found that DAR increased with increasing stroke severity, particularly in the moderate and severe groups. There was a significant difference between DAR of those with large strokes and both controls and those in the small stroke category (p < 0.01, F = 5.79, df = 3, 42, partial η^2^ = 0.293). We saw similar results when we calculated the DTABR (p < 0.01, F = 4.29, df = 3, 42, partial η^2^ = 0.234), as seen in Figure 2D. In both DAR and DTABR, there was an increased power in the ipsilateral side of the brain for patients in the large stroke category. This effect was insignificant; however, an interesting effect to note as the other severities and controls showed the opposite effect.

We analysed the general spectra using fooof to determine if the band ratio effects were due to non-oscillatory changes in brain activity (Figure 3). Specifically, the slope of the background spectra did not change between severities (p = 0.6336, F = 0.58, df = 3, 21). The intercepts of the of the background spectra also had no changes between severities (p = 0.7743, F = 0.37, df = 3, 21).

**Figure 3.**
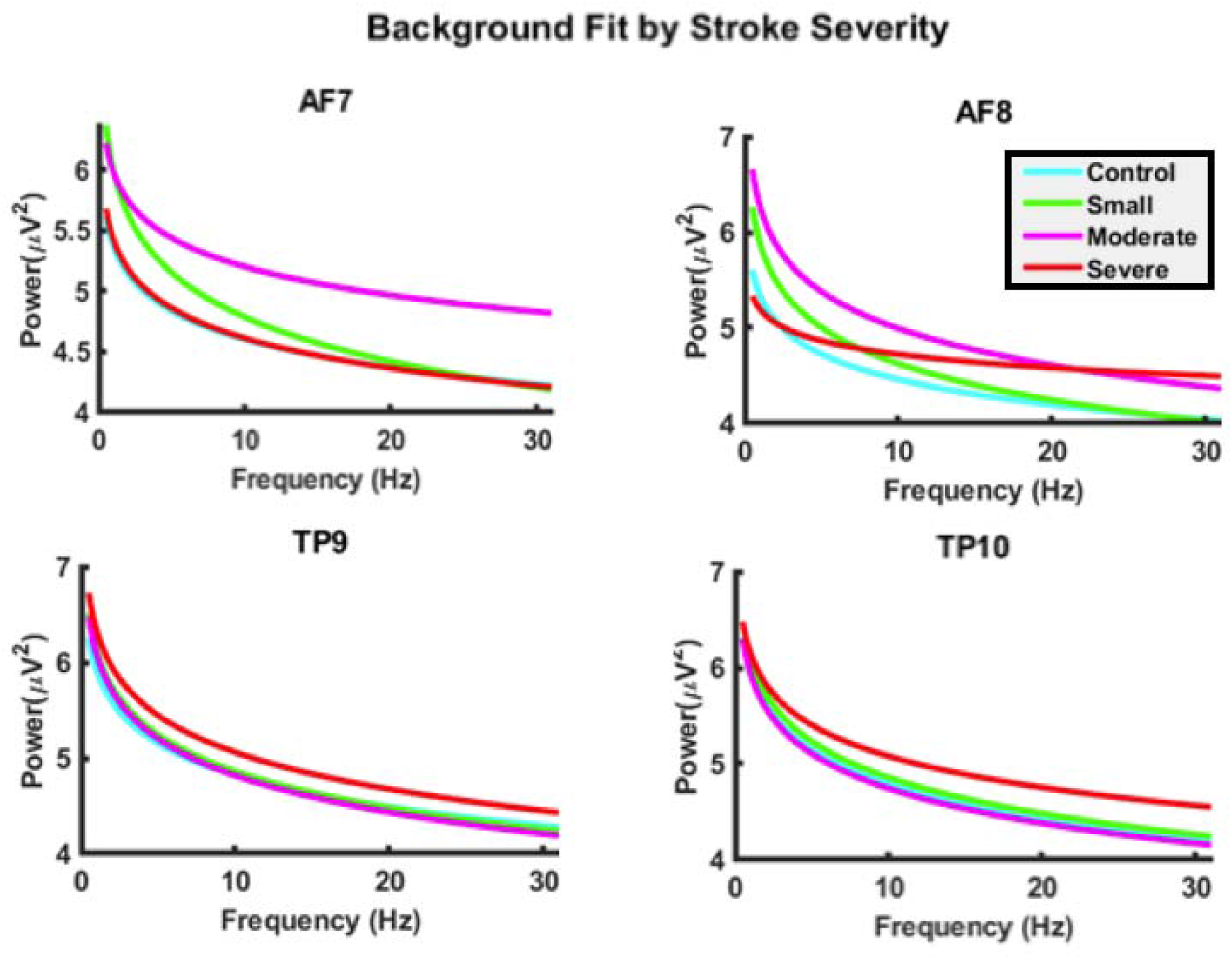
Background fit of data from fooof analysis at each electrode by severity. Non-oscillatory activity was not different between severities, indicating that band-ratios are not affected by background brain activity levels.

The RMS of the accelerometer data found a significant increase in movement in the Z-plane when comparing large strokes to small strokes and controls (p ≈ 0, F = 7.13, df = 6, 63 partial η^2^ = 0.404). There was no effect present in the Y-plane and a significant decreasing effect present in the X-plane when comparing large strokes to small strokes and controls (Figure 4). Data from the gyroscope had no significant differences; however, the variability in movement is more consistent among controls when compared to the other groups, in all planes.

**Figure 4.**
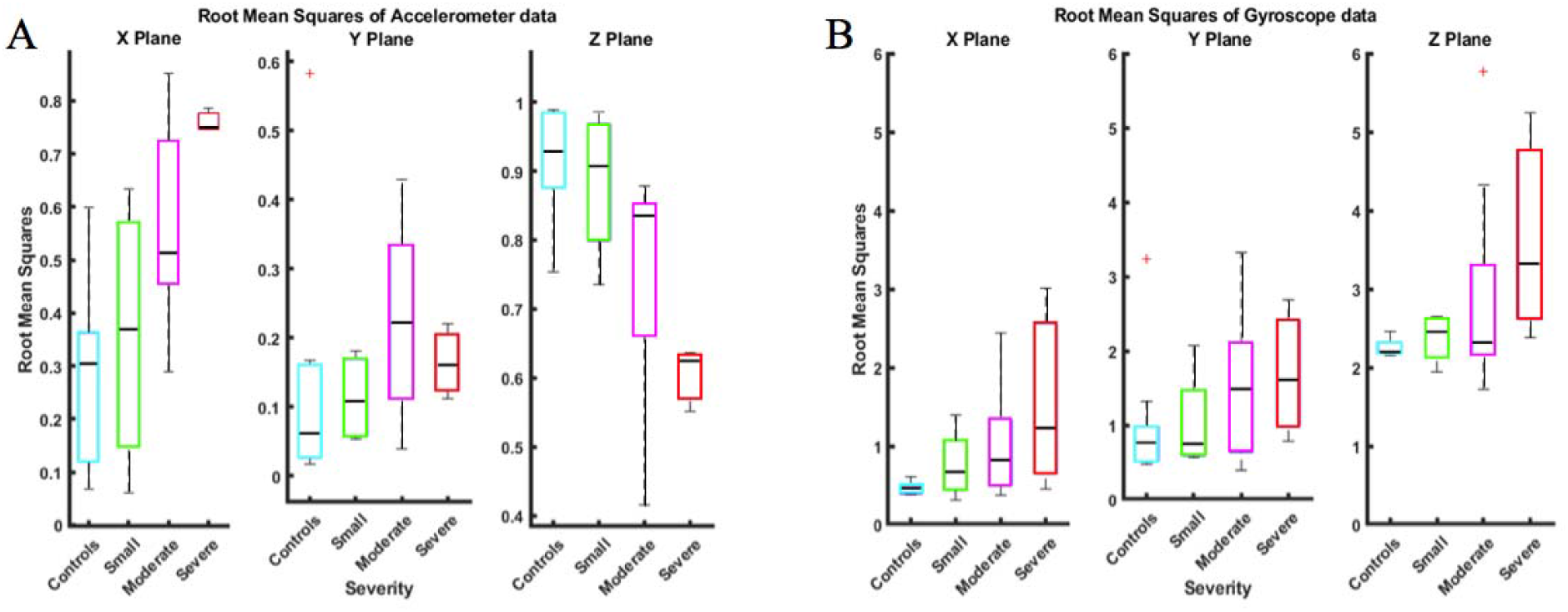
RMS for accelerometer (A) and gyroscope (B) across three planes of movement. RMS of accelerometer data increased with increasing stroke severity in the X-plane and decreased with stroke severity in the Z-plane. No significant group differences in the gyroscope data. Data are presented as mean ± SEM.

The severity classification has an accuracy of 0.36 and sensitivity for controls, small strokes, moderate strokes, and large strokes, of 0.67, 0, 0.33, and 0, respectively, as seen in Figure 5A. The classification tree that groups by stroke or control has an accuracy of 0.67, with a sensitivity and specificity of 0.75 and 0.33, respectively (Figure 5B). The final classification tree groups by higher-risk strokes or small strokes and controls has an accuracy of 0.76, with a sensitivity and specificity of 0.63 and 0.86, respectively (Figure 5C).

**Figure 5.**
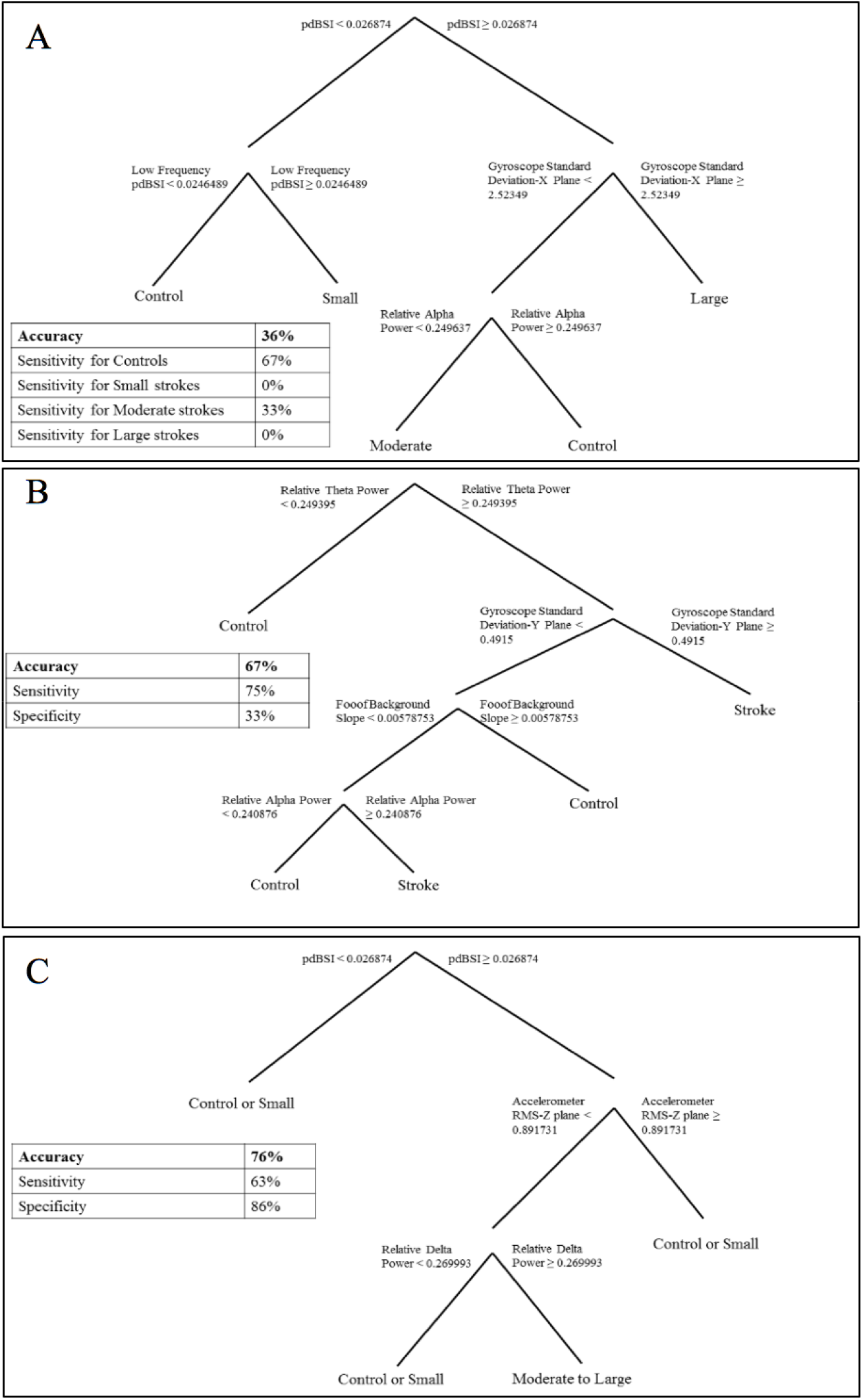
Classification trees for determining (A) stroke severity, (B) presence of stroke, (C) higher-risk stroke. Predicting stroke severity was the least accurate model and predicting higher-risk strokes was the most accurate model.

## Discussion

This pilot study indicates that stroke and stroke severity can be detected with the Muse™ EEG system. Brain symmetry differs between stroke patients and controls at certain frequencies. Additionally, DAR and DTABR are increased in moderate and severe strokes, indicating that there is a slowing in brain activity. Further, the Muse™ set up took approximately 5 minutes and was tolerated even by patients with more severe deficits, making this system feasible for future use in an ambulance. The short set-up time, combined with simple qEEG measures, makes this system a promising method for distinguishing strokes from stroke mimics and identifying those strokes associated with LVO that need to be triaged preferentially to comprehensive stroke centres with endovascular thrombectomy capability.

Although many papers find increased asymmetry after stroke,^7,10,32^ it is difficult to compare findings directly. Most studies that assess brain symmetry compare average power, rather than power across frequencies, as we have done here. This ulterior method may mask differences in pdBSI across frequencies, such as our observed increased asymmetry at lower frequencies and decreased asymmetry at higher frequencies. Further, many researchers compute pdBSI from 1-25 Hz, whereas we found the most pronounced differences in pdBSI from ~20-30 Hz, which may be driving the differences seen. A recent pilot study found, using the Muse™, that stroke patients had increased brain asymmetry compared to controls using the revised brain symmetry index.^27,33^ This is contrary to our findings, which show overall decreased asymmetry in the stroke patients. The revised BSI compares the symmetry at each frequency across the electrodes, whereas the pdBSI considers the symmetry between each homologous pair of electrodes at each frequency.^10^ In this case, we compared symmetry using one homologous pair of electrodes, so this would not affect these results, but may affect comparisons to other studies of brain symmetry after stroke. However, Gottlibe et al. filtered their data from 0.16-76 Hz and therefore may be picking up on muscle artifacts in the higher frequency range, which may be the reason for differences between these findings and our findings.^34^ In this study, we show that reducing brain symmetry measures to a single number may be masking important information, such as the symmetry differences between lower and higher frequencies.

Here, we found an increase in DAR and DTABR in moderate and severe strokes. This increase indicates a slowing in brain activity due to ischemia and likely impaired CBF, similar to the findings of other studies in acute stroke.^9,35^ Interestingly, we did not see a significant increase in either DAR or DTABR in small strokes, indicating that this measure may be useful in distinguishing strokes by severity and identify those patients most likely to have LVO. This effect of stroke severity on DAR is supported by studies that show DAR predicts outcome and relates to stroke recovery.^10,18^

With the Muse™, we can also look at participant movement using data obtained from the accelerometer and the gyroscope during recording. Here, we found that stroke patients had more variability in acceleration in the X-axis and less in the Z-axis, which increased with increasing severity. This finding equates to more movement in tilting the head from left to right (as in, tilting ear to shoulder), and less movement shaking the head from left to right (as in, “shaking your head no”). These findings may indicate movement differences due to motor system impairments or attentional differences due to contralateral neglect. However, since data was collected with eyes closed, we believe differences are likely due to impaired motor function, including postural support. Interestingly, there were no differences in the movement data from the gyroscope, which can also measure rotation. This indicates that the accelerometer may be a more useful tool for detecting movement differences between stroke patients and controls, and that differences in general movement may be more predictive than rotational differences.

In the literature, there are multiple qualitative and quantitative aspects of EEG that may differentiate stroke patients from controls. Here, we used decision trees to determine which of the quantitative EEG and movement measures we looked at were the most useful in determining which participants had a stroke and to which severity category they belonged. We used the TreeBagger function in MATLAB, which controls for overfitting to the sample population by creating bootstrap-aggregated decision trees. Thus, although we have “wide data,” with many predictor variables and a small sample size, this method mitigates some of the risks of overfitting. Here, we found that our model for determining severity had low accuracy and had difficulty (0% sensitivity) at classifying small strokes and large strokes. However, model accuracy improved when data was used to determine if participants were in the stroke group (any size) or control group. This model had 67% accuracy, 75% sensitivity, and 33% specificity. Features included in this tree were: relative theta and alpha power, variability in the Y-axis using the gyroscope, and the slope of the background spectra (the aperiodic component of the EEG spectra). The model improved further when classifying higher-risk moderate and large strokes from other patients (i.e., those that would be more likely to need to get to a stroke centre vs. a standard hospital). This model had 76% accuracy, 63% sensitivity, and 86% specificity. Important predictors in this model were: brain symmetry, variability in the Z-axis using the accelerometer, and relative delta power. These decision trees provide a simple way to predict stroke incidence and stroke severity using multiple measures from the Muse™ instead of relying on one index.

Our pilot study has notable limitations. First, we assessed patients an average of 3.71 days (range: 0-16 days) after stroke onset. In the subacute phase of stroke, as we have assessed in this study, reperfusion and restoration of CBF likely change the EEG signatures as compared to the hyper-acute phase of stroke, when CBF is severely decreased. Indeed, different levels of CBF are associated with different EEG frequency alterations.^8^ However, assessing patients in the subacute phase was more practical for this study as the goal was to establish the initial feasibility of this stroke detection method. Second, we have a small sample size and are likely underpowered to detect effects. When analyzing the data by stroke severity, hemisphere, location of stroke, etc., the subgroups have a small sample size that may not be representative of that population. Therefore, our follow up study will validate these findings using a much larger sample size so that subgroup analyses will be adequately powered. Third, scalp measures of EEG measure cortical activity, and the effects of stroke on cortical activity likely vary depending on stroke location.^12^ In this sample, only three stroke patients had subcortical stroke, leaving us underpowered to detect effects of stroke location on EEG. However, when using classification trees to predict stroke location, none of the EEG measures significantly predicted stroke location (model R^2^=0.000171), suggesting that stroke location had minor effects on EEG measures and that EEG disturbances caused by stroke may be largely global. Finally, prehospital stroke scale scores were not available for study patients. As a result, it remains unclear to what extent EEG will improve the diagnostic accuracy above that of currently used EMS stroke scales.

Our initial results suggest that qEEG measures, such as pdBSI, DAR, DTABR, and head movement variability may be able to distinguish stroke severities when measured using the Muse™. However, these findings will need to be validated in a larger sample and at an earlier time after stroke onset, as brain activity changes with the progression of stroke injury. Future work will assess patients earlier, such as in the emergency department or in the ambulance, to determine the utility of using EEG in the hyper-acute phase of stroke. Further studies will help elucidate the feasibility and utility of using the Muse™ as an inexpensive and early detection tool to use in ambulances to aid in the diagnosis of stroke and the detection of stroke from LVO.

## Data Availability

Anonymized data will be shared upon request from qualified investigators

## Acknowledgements

The authors thank Paige Fairall for assistance in patient recruitment and data collection. CMW is supported by a Canadian Institutes of Health Research graduate scholarship. BHB is supported by Alberta Innovates Health Solutions Collaborative Research and Innovations Opportunities and the Partnership for Research and Innovation in the Health System Grants. This research was supported by start-up funds from the Faculty of Science at the University of Alberta and an NSERC Discovery Grant (#RES0024267) awarded to KEM.

## Appendix

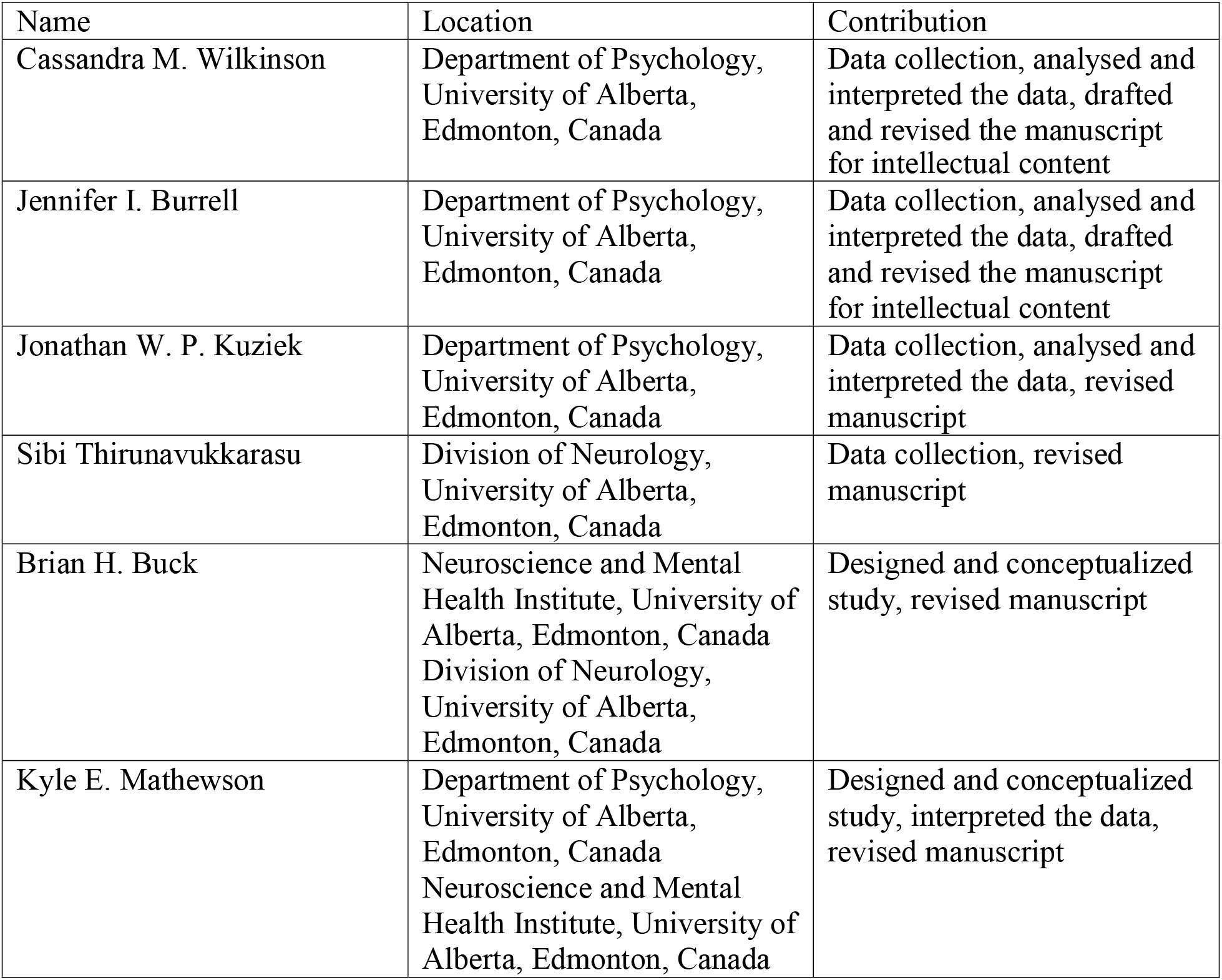

## References

1. Johansen HL, Wielgosz AT, Nguyen K, et al. Incidence, comorbidity, case fatality and readmission of hospitalized stroke patients in Canada. Can J Cardiol. Epub ahead of print 2006. DOI: 10.1016/S0828-282X(06)70242-2.

2. Walker GB, Zhelev Z, Henschke N, et al. Prehospital stroke scales as screening tools for early identification of stroke and transient ischemic attack. Stroke. Epub ahead of print 2019. DOI: 10.1161/STROKEAHA.119.026527.

3. Brandler ES, Sharma M, Sinert RH, et al. Prehospital stroke scales in urban environments: A systematic review. Neurology. Epub ahead of print 2014. DOI: 10.1212/WNL.0000000000000523.

4. Smith EE, Kent DM, Bulsara KR, et al. Accuracy of Prediction Instruments for Diagnosing Large Vessel Occlusion in Individuals With Suspected Stroke: A Systematic Review for the 2018 Guidelines for the Early Management of Patients With Acute Ischemic Stroke. Stroke. Epub ahead of print 2018. DOI: 10.1161/STR.0000000000000160.

5. Walsh KB. Non-invasive sensor technology for prehospital stroke diagnosis: Current status and future directions. International Journal of Stroke. Epub ahead of print 2019. DOI: 10.1177/1747493019866621.

6. Schneider AL, Jordan KG. Regional Attenuation WithOut Delta (RAWOD): A distinctive EEG pattern that can aid in the diagnosis and management of severe acute ischemic stroke. Neurodiagnostic Journal. Epub ahead of print 2005. DOI: 10.1080/1086508x.2005.11079517.

7. Agius Anastasi A, Falzon O, Camilleri K, et al. Brain symmetry index in healthy and stroke patients for assessment and prognosis. Stroke Res Treat. Epub ahead of print 2017. DOI: 10.1155/2017/8276136.

8. Jordan KG. Emergency EEG and continuous EEG monitoring in acute ischemic stroke. Journal of Clinical Neurophysiology. Epub ahead of print 2004. DOI: 10.1097/01.WNP.0000145005.59766.D2.

9. Finnigan S, Wong A, Read S. Defining abnormal slow EEG activity in acute ischaemic stroke: Delta/alpha ratio as an optimal QEEG index. Clin Neurophysiol. Epub ahead of print 2016. DOI: 10.1016/j.clinph.2015.07.014.

10. Sheorajpanday RVA, Nagels G, Weeren AJTM, et al. Quantitative EEG in ischemic stroke: Correlation with functional status after 6months. Clin Neurophysiol. Epub ahead of print 2011. DOI: 10.1016/j.clinph.2010.07.028.

11. Giaquinto S, Cobianchi A, Macera F, et al. EEG Recordings in the course of recovery from stroke. Stroke. Epub ahead of print 1994. DOI: 10.1161/01.STR.25.11.2204.

12. Fanciullacci C, Bertolucci F, Lamola G, et al. Delta power is higher and more symmetrical in ischemic stroke patients with cortical involvement. Front Hum Neurosci. Epub ahead of print 2017. DOI: 10.3389/fnhum.2017.00385.

13. Macdonell RAL, Donnan GA, Bladin PF, et al. The Electroencephalogram and Acute Ischemic Stroke: Distinguishing Cortical From Lacunar Infarction. Arch Neurol. Epub ahead of print 1988. DOI: 10.1001/archneur.1988.00520290048013.

14. Doerrfuss JI, Kilic T, Ahmadi M, et al. Quantitative and Qualitative EEG as a Prediction Tool for Outcome and Complications in Acute Stroke Patients. Clinical EEG and Neuroscience. Epub ahead of print 2019. DOI: 10.1177/1550059419875916.

15. Aminov A, Rogers JM, Johnstone SJ, et al. Acute single channel EEG predictors of cognitive function after stroke. PLoS One. Epub ahead of print 2017. DOI: 10.1371/journal.pone.0185841.

16. Schleiger E, Sheikh N, Rowland T, et al. Frontal EEG delta/alpha ratio and screening for post-stroke cognitive deficits: The power of four electrodes. Int J Psychophysiol. Epub ahead of print 2014. DOI: 10.1016/j.ijpsycho.2014.06.012.

17. Finnigan SP, Walsh M, Rose SE, et al. Quantitative EEG indices of sub-acute ischaemic stroke correlate with clinical outcomes. Clin Neurophysiol. Epub ahead of print 2007. DOI: 10.1016/j.clinph.2007.07.021.

18. Leon-Carrion J, Martin-Rodriguez JF, Damas-Lopez J, et al. Delta-alpha ratio correlates with level of recovery after neurorehabilitation in patients with acquired brain injury. Clin Neurophysiol. Epub ahead of print 2009. DOI: 10.1016/j.clinph.2009.01.021.

19. Liew SL, Garrison KA, Ito KL, et al. Laterality of poststroke cortical motor activity during action observation is related to hemispheric dominance. Neural Plast. Epub ahead of print 2018. DOI: 10.1155/2018/3524960.

20. Van Putten MJAM, Peters JM, Mulder SM, et al. A brain symmetry index (BSI) for online EEG monitoring in carotid endarterectomy. Clin Neurophysiol. Epub ahead of print 2004. DOI: 10.1016/j.clinph.2003.12.002.

21. Xin X, Gao Y, Zhang H, et al. Correlation of continuous electroencephalogram with clinical assessment scores in acute stroke patients. Neurosci Bull. Epub ahead of print 2012. DOI: 10.1007/s12264-012-1265-z.

22. Zhang SJ, Ke Z, Li L, et al. EEG patterns from acute to chronic stroke phases in focal cerebral ischemic rats: Correlations with functional recovery. Physiol Meas. Epub ahead of print 2013. DOI: 10.1088/0967-3334/34/4/423.

23. Lu XCM, Williams AJ, Tortella FC. Quantitative electroencephalography spectral analysis and topographic mapping in a rat model of middle cerebral artery occlusion. Neuropathol Appl Neurobiol. Epub ahead of print 2001. DOI: 10.1046/j.1365-2990.2001.00357.x.

24. Krigolson OE, Williams CC, Norton A, et al. Choosing MUSE: Validation of a low-cost, portable EEG system for ERP research. Front Neurosci. Epub ahead of print 2017. DOI: 10.3389/fnins.2017.00109.

25. Ratti E, Waninger S, Berka C, et al. Comparison of medical and consumer wireless EEG systems for use in clinical trials. Front Hum Neurosci. Epub ahead of print 2017. DOI: 10.3389/fnhum.2017.00398.

26. Titgemeyer Y, Surges R, Altenmüller DM, et al. Can commercially available wearable EEG devices be used for diagnostic purposes? An explorative pilot study. Epilepsy Behav. Epub ahead of print 2019. DOI: 10.1016/j.yebeh.2019.106507.

27. Gottlibe M, Rosen O, Weller B, et al. Stroke identification using a portable EEG device - A pilot study. Neurophysiol Clin. Epub ahead of print 2020. DOI: 10.1016/j.neucli.2019.12.004.

28. Barachant A, Morrison D, Banville H, et al. Muse-LSL. Epub ahead of print 2019. DOI: 10.5281/zenodo.3228861.

29. Caplan JB, Madsen JR, Raghavachari S, et al. Distinct patterns of brain oscillations underlie two basic parameters of human maze learning. J Neurophysiol. Epub ahead of print 2001. DOI: 10.1152/jn.2001.86.1.368.

30. Donoghue T, Dominguez J, Voytek B. Electrophysiological Frequency Band Ratio Measures Conflate Periodic and Aperiodic Neural Activity. *bioRxiv Prepr* 2020; 1-33.

31. Zhang J, Lin F, Xiong P, et al. Automated detection and localization of myocardial infarction with staked sparse autoencoder and treebagger. IEEE Access. Epub ahead of print 2019. DOI: 10.1109/ACCESS.2019.2919068.

32. Van Putten MJAM, Tavy DLJ. Continuous quantitative EEG monitoring in hemispheric stroke patients using the brain symmetry index. In: Stroke. 2004. Epub ahead of print 2004. DOI: 10.1161/01.STR.0000144649.49861.1d.

33. van Putten MJAM. The revised brain symmetry index. Clin Neurophysiol. Epub ahead of print 2007. DOI: 10.1016/j.clinph.2007.07.019.

34. Muthukumaraswamy SD. High-frequency brain activity and muscle artifacts in MEG/EEG: A review and recommendations. Frontiers in Human Neuroscience. Epub ahead of print 2013. DOI: 10.3389/fnhum.2013.00138.

35. Bentes C, Peralta AR, Viana P, et al. Quantitative EEG and functional outcome following acute ischemic stroke. Clin Neurophysiol. Epub ahead of print 2018. DOI: 10.1016/j.clinph.2018.05.021.

